# Interactive Psychometrics for Autism with the Human Dynamic Clamp: Interpersonal Synchrony from Sensory-motor to Socio-cognitive Domains

**DOI:** 10.1101/19013771

**Authors:** F. Baillin, A. Lefebvre, A. Pedoux, Y. Beauxis, D. Engemann, A. Maruani, F. Amsellem, T. Bourgeron, R. Delorme, G. Dumas

## Abstract

**Background:** The Human Dynamic Clamp (HDC) is a human-machine interface for studying realistic social interaction under controlled and reproducible conditions. Here, we propose to probe the validity of the HDC as psychometric instrument for quantifying social abilities in children with Autism Spectrum Disorder (ASDs) and neurotypical development.

**Methods:** To study behavioral synchrony, we derived from interaction with the HDC avatar, five standardized scores following a gradient from sensory-motor and motor to higher socio-cognitive skills, in a sample of 155 individuals (113 with ASDs, 42 typically developing participants; aged 5 to 25 years; IQ>70).

**Results:** We conducted regression analyses with normative modeling on global scores according to four sub-conditions (avatar behavior “cooperative/competitive”, human task “in-phase/anti-phase”, diagnosis and age at inclusion). Children with ASDs tend to have significant lower scores than controls for motor skills. Independently of the phenotype, socio-cognitive skills increase with developmental age, while being affected by the ongoing task and the behavior of the avatar.

**Discussion:** The weaker performance in participants with ASDs for motor skills suggests convergent validity for this score of the HDC during social interaction. Results provide additional evidence of a relationship between sensory-motor and socio-cognitive skills. As we found a significant main effect of age at inclusion, HDC may be used as a marker of aging of socio-cognitive skills during real-time social interaction.

**Conclusion:** Through its standardized and objective evaluation, the HDC not only represents a valid paradigm for the study of interpersonal synchrony but also a clinically relevant psychometric instrument for the evaluation and stratification of socio-motor dysfunctions.

## 1. INTRODUCTION

Autism Spectrum Disorders (ASDs) are a family of complex neurodevelopmental disorders American Psychiatric Association (APA) (2013) which are defined by the co-occurrence of significant differences in the development of social communication and interaction, and the restricted and repetitive nature of behaviors and interests. The prevalence of these disorders has increased in recent years from less than 1 in 1000 individuals to 1 in 58 (Baxter et al. 2015; Fombonne 2003). The strong heterogeneity of the conditions complicates the development of psychometric assessment tools that allow for a personalized and thorough evaluation of a child’ s skills (Vivanti et al. 2014). Indeed, ASDs are highly diverse at both phenotypical (Georgiades et al. 2013) and genetic levels (Huguet, Ey, et Bourgeron 2013), with more than 50% of the patients suffering from at least four other psychiatric comorbid conditions (Soke et al. 2018). The major societal challenge of improving early diagnosis thus requires the discovery of robust and scalable biomarkers related to social communication dysfunction in humans.

An important key for increasing our comprehension and early detection of ASD-specific social dysfunction may lie on the Interpersonal Synchrony (IS). IS can be defined as a rhythmic matching of actions in time and in phase with another person, based on non-verbal behaviors (Mogan et al. 2017). IS comprises multiple components, such as socio-cognitive, sensory-motor, and motor skills, as well as adaptive capacities (Nebel et al. 2016; Xavier et al. 2017). At the behavioral level, IS can be measured through micro-level detection of bonding-related behaviors (Atzil et al. 2011), through frame-by-frame analysis of video (Dumas et al. 2010), or even machine learning (Delaherche, Dumas, Chetouani, et Cohen 2012).

The Human Dynamic Clamp (HDC) is a new paradigm of human-machine interaction for the study of neurobehavioural processes involved in IS (Dumas et al. 2014; Kelso et al. 2009). It allows to recreate a dynamic bidirectional interaction in real time between a human and a virtual avatar, controlled by empirical grounded models. The HDC paradigm has already been validated in an adult with a neurotypical development through the study of underlying neural processes linking sensory-motor and socio-cognitive processes (Dumas et al. 2019), induction of emotional reaction (Zhang et al. 2016), and in a Virtual Teacher (VT) configuration of HDC which allows to change the behavioral repertoire by internalizing new interpersonal coordination patterns, thus opening HDC to rehabilitation (Kostrubiec et al. 2015).

IS seems to be substantively impaired in children and adolescents with ASDs (Xavier et al. 2018). A few studies among children (6-11y) (Romero et al. 2018) and adolescents (10-16.5y) (Noel et al. 2018) have explored IS in automated motion analysis to quantify movements of body-parts. Still, the exploratory paradigms are mainly rhythmic in children with ASDs (3.5-10y) (Marsh et al. 2013; Kaur et al. 2018; Fitzpatrick et al. 2017a; 2017b) and in adolescents (12-17y) (de Marchena et Eigsti 2010; Fitzpatrick et al. 2016). However, even if children with ASDs face difficulties at coordinating their body during a social exchange, social embodiment seems preserved and correlates to social cognitive ability (Romero et al. 2018).

One hypothesis currently under investigation suggests that motor and sensory-motor skills development are linked to social cognition and cognitive development (Bhat et al. 2016; Kaur et al., 2018). ASDs are frequently found to be associated with difficulties to attribute mental states to oneself and to others (Zalla et Korman 2018), where intention attribution is characterized by an appraisal based on intention underlying someone else’ s action (Hilton et Kuhlmeier 2019). In addition to primary dysfunctions in social communication skills, deficits in perceptual motor performance are found in between 50% to 80% of children diagnosed with ASDs (moving with awareness, integrated self, proprioceptive feedback, visuo-perceptual performance, sensory integration) (Kaur et al. 2018; Bhat et al. 2011; Torres et al. 2013; Righi et al. 2018; Greenfield et al. 2015; Noel et al. 2018; Lim et al. 2017). About 80% of these patients also show impairments in motor skills such as praxis, basic motor control, postural control, gait abnormalities, motor coordination, manual dexterity, gross and fine motor skills, gestures in complex movement sequences (Fournier et al. 2010; Kaur et al. 2018; Xavier et al. 2018).

Interventions targeting the development of IS is promising and shows evidence for plasticity (Landa et al. 2011; Zhou et al. 2018; Franz et Dawson 2019). Early intervention and detection that directly focus its development showed preliminary evidence of potent effects on communication and motor skills (Eapen et al. 2013) or on later language and social abilities (Landa et al. 2011).

Indeed, language, cognitive ability, social engagement and motor skills have emerged as the most robust predictors of ASD among toddlers (Landa et al. 2013; Ozonoff et al. 2018; Iverson et al. 2019) and during childhood and adolescence (Stahmer et al. 2011). Thus, an early dysfunction in IS could have cascading consequences and even participate in explaining the heterogeneity in ASDs. This observation reflects the difficulty of assessments by reliable and scalable markers of IS over several ages and the need for a personalized analysis.

In this work, we first want to validate in children with neurotypical development how the HDC can measure different behavioural mechanisms involved in social dynamic interactions, and then to evaluate how the HDC can assess the alteration of IS in ASDs. The secondary objective has required to standardize the test and develop indicators that measure and identify socio-cognitive and sensory-motor markers. Finally, to take into account the heterogeneity of ASDs, we also integrated the developmental trajectories with normative modeling and tried to test how the HDC behavioral measures can provide reproducible and reliable clinical markers.

## 2. METHODS

### Sample

We enrolled in the study a sample of 155 individuals composed of 113 participants with ASDs and 42 participants with typical development (Table 1). They were all recruited at the Child Psychiatry Department of the Robert Debré Universitary Hospital, Paris (France).

**Table 1.**
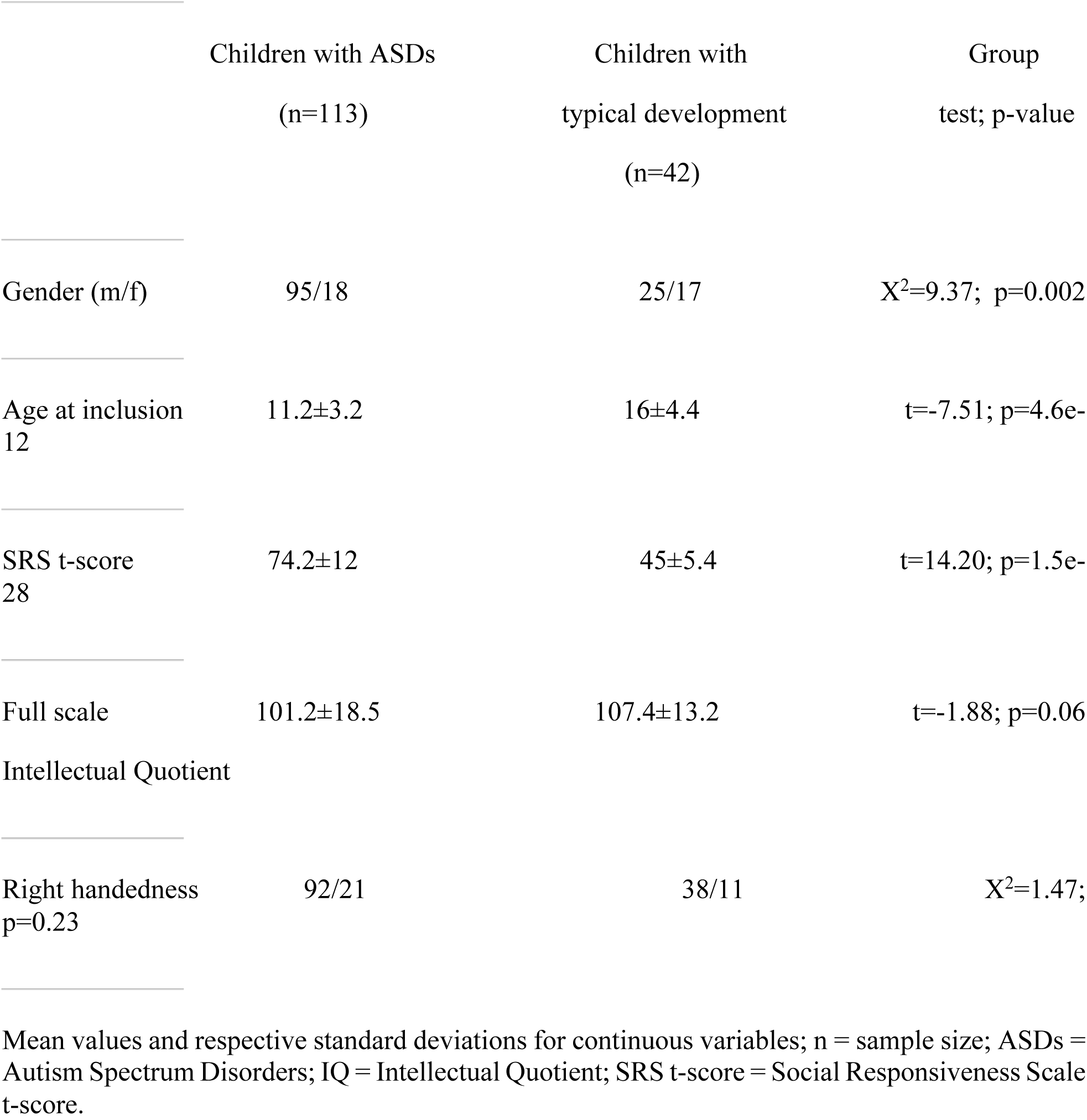
Demographic and clinical characteristics of the subjects enrolled in the study.

Patients with ASDs were included after a systematic clinical and medical examination including negative blood tests results for Fragile-X and exclusion of participants carrying a large deletion over 2 Mb detected by the Illumina 700 SNPs array. Final diagnosis of ASD was based on DSM-5 criteria and the information outcomes from the Autism Diagnostic Observation Schedule-Second Edition (ADOS-II) (Lord C et al. 2012), the Autism Diagnostic Interview-Revised (ADI-R) (Lord, Rutter, et Le Couteur 1994), the Social Responsiveness Scale - 2nd Edition (SRS-2) (Constantino 2013) for the dimensional diagnosis of social skills and data from experts in the field. Intellectual functioning for all participants was estimated with the Wechsler Intelligence Scale for Children and Adolescents - 5th Edition (WISC-V). Participants with neurotypical development were from the general population and reported no personal or familial history of ASDs or axis I psychiatric conditions requiring specific needs.

An assessment of dexterity and motor coordination of hands and fingers was made with the Purdue Pegboard. For this study, we used the versions with charts defined on a population aged 5 to 15 years 11 months and beyond the age of 16 (Gardner et Broman 1979; Yeudall et al. 1986). Only the “preferred hand score”: place the most stems using the preferred hand on a row in thirty seconds was conserved. A z-score is calibrated according to age and gender. Children were also assessed with the Child Neuropsychological Assessment-Second Edition (NEPSY-II) (Kemp et Korkman 2010) to specifically explore Affect Recognition (AF) and Theory of Mind (TOM).

This study was carried out in accordance with the recommendations of the local ethics committee of Hospital Robert Debré. All the parents of the participants gave written informed consent in accordance with the Declaration of Helsinki. The protocol was approved by the Inserm Ethics Committee (study approval no. 08-029).

### Human Dynamic Clamp (HDC) paradigm

The HDC system (Kostrubiec et al. 2015; Dumas et al. 2014) is a human-machine interface. It consists of three parts: a sensor measuring the movement of the participant’ s index which is positioned in the moving sensor a mathematical model integrating the position and speed of the human participant to simulate in real time the behaviour of an avatar; and a screen facing the participant where the actions of the Virtual Partner (VP) appear as an index hand. The HDC software computes in real time the corresponding position of the VP (Figure 1). Three main positions for the indexes are possible: extension, in the axis of the wrist, flexion. At the beginning of each trial, an instruction was given to the participant to synchronize its movement in-phase i.e., to synchronized her/his movements to those of the partner or in anti-phase meaning that s/he has to synchronize her/his movements with a half-period offset between the phase of the partner and the participant. In this experiment, all participants were instructed that the partner will be half of the time virtual (i.e., movements will be computer driven) and half of the time a real sex-, age-matched human performing the same task in another room of the hospital (although in this case, the partner was also virtual). The protocol was composed of 40 trials, divided into four blocks. The instructions to the participant stayed the same within each block. The instruction for the first block was randomly assigned at the beginning of the experiment. During the trials, the VP could adopt a “cooperative” or “competitive” behavior, meaning that its shares the same goal as the participant or has the opposite goal to the one assigned to the participant (i.e., VP aims at in-phase coordination when participant aims at anti-phase; and vice versa). Behavior of the VP was randomized across all trials, disregarding block structure.At the end of each trial, we asked the participant if s/he felt like s/he was playing with a human or a VP, and if s/he could quantify the level of cooperativeness to competitiveness of the partner.

**Figure 1.**
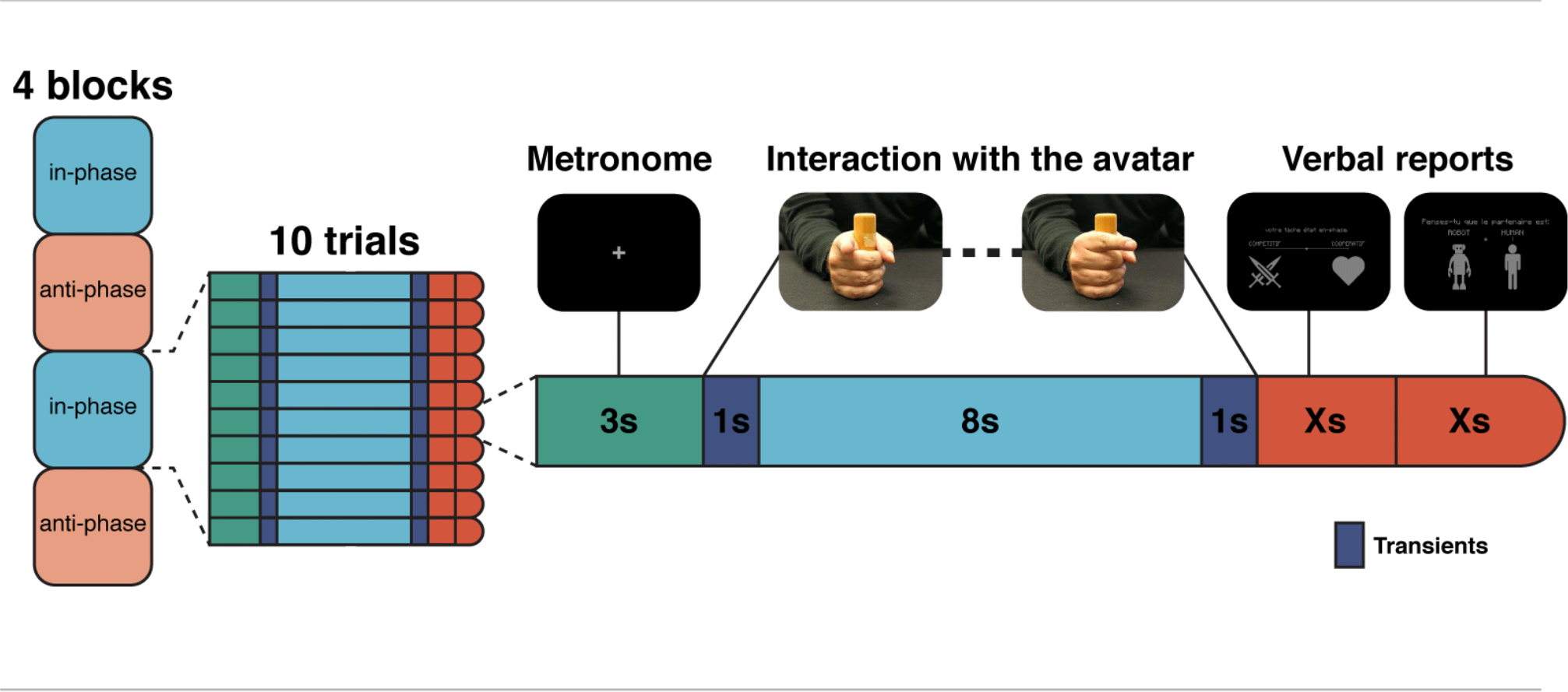
Experimental design. Structure of the protocol with four blocks alternating “in-phase/anti-phase”. Each block is divided into ten trials (Left). Each trial starts with a phase while the participant has to synchronize with the sound of a metronome for 3 seconds. Then, the participant interacts with the avatar according to the instruction (i.e. here, “in-phase”). At the end of the trial, a report of two questions is made directly on the screen. First of all, the impression of the participant on the competitive or cooperative behavior of the avatar and his/er impression on the human nature of the avatar (Right). We represented screenshots of what participants could see (Top).

**Figure 2.**
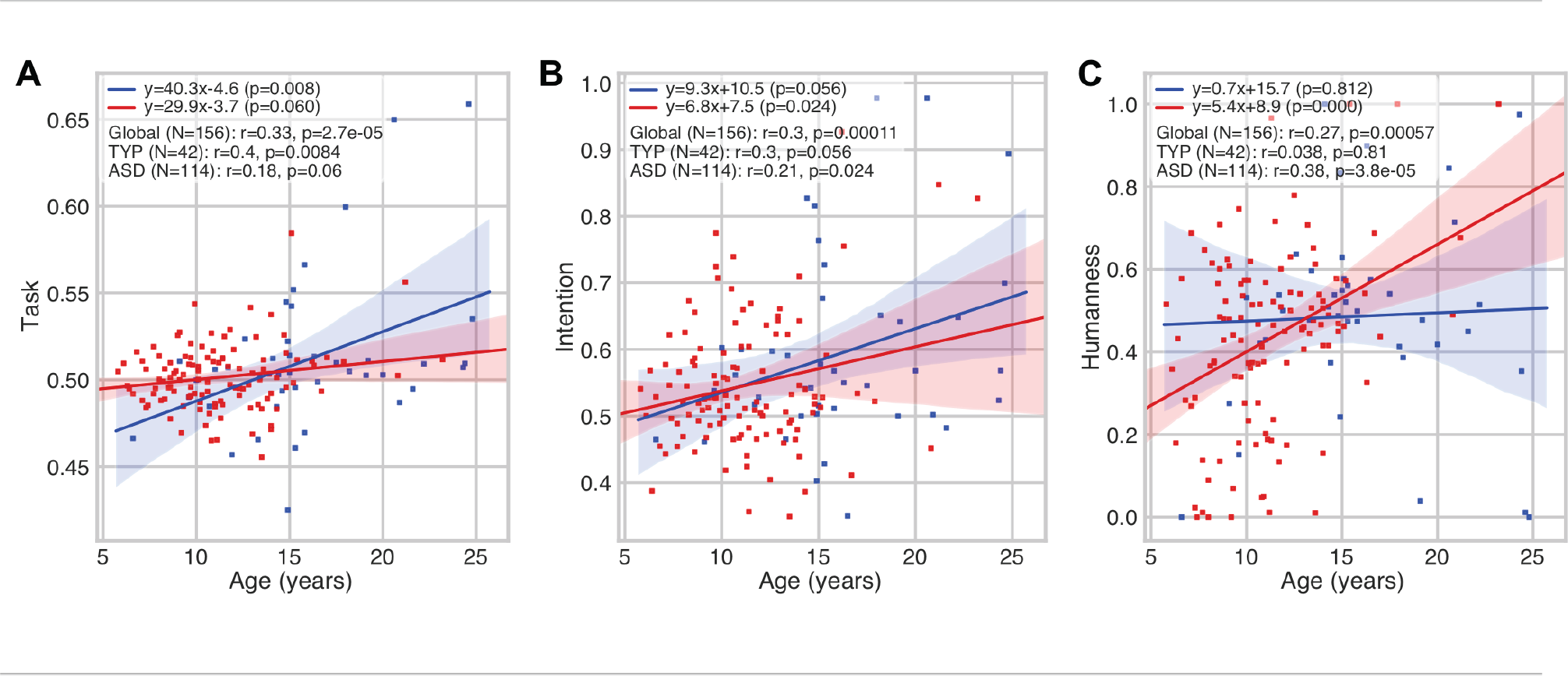
Developmental aspect of higher level of IS. Correlations between age at inclusion and task comprehension (A), intention attribution (B) and humanness (C) scores. These three scores show remarkable and positive correlation with age, suggesting the developmental aspect of socio-cognitive skills.

**Figure 3.**
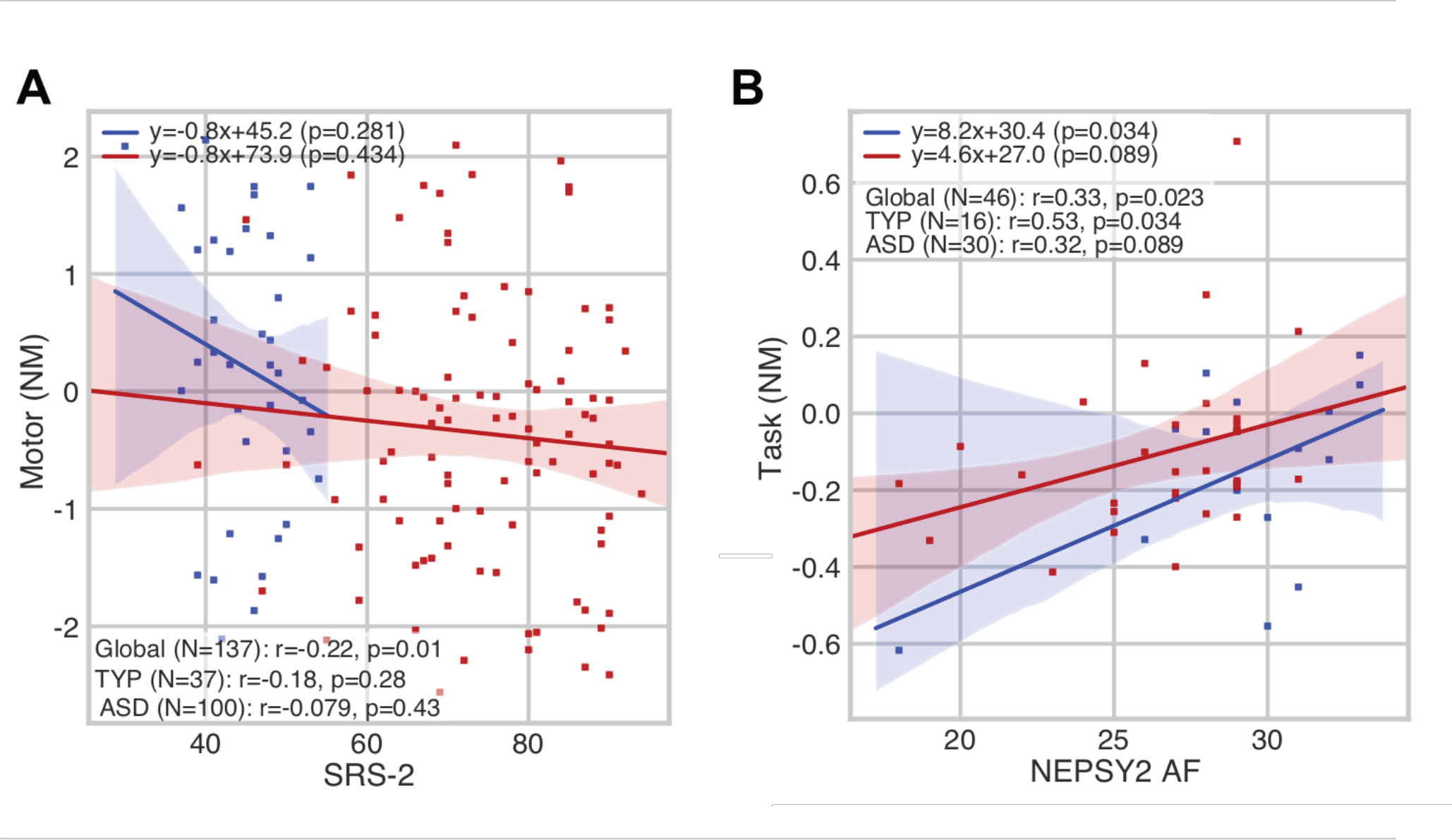
Significant correlations between socio-cognitive and motor skills in children with (in red) autism Spectrum Disorder (ASD)s or with typical development (TYP) (in blue): A) SRS-2 vs Motor score: A dimensional diagnosis of ASD correlates with lower levels of motor skills; and B) NEPSY-II Affect Recognition (AF) vs. HDC Task score: Greater cognitive abilities correlates with a higher levels of affect recognition skills; NM = Normative Models

### HDC behavioral measures

In our study, five scores (between 0 and 1, 0 being the worst) of the HDC paradigm were automatically generated from sensory-motor to representational dimensions of social cognition:

i/ a motor score: which measure the difference of amplitude of imitative gestures between the participant and the VP. A mean is obtained for each trial.

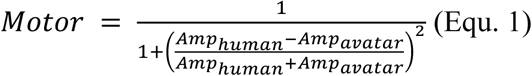

where *Amp*_*human*_and *Amp*_*avatar*_are respectively the amplitude of the movement of the participant and the virtual avatar, both extracted from a Hilbert transform;

ii/ a coordination score which corresponds to the temporal index of imitation. This measure is based on the frequency parameter and evaluate the shift of frequency between that of the model and that of the participant. A mean is obtained for each trial.

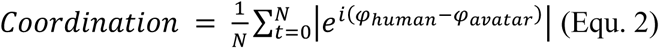

where *φ*_*human*_ and *φ*_*avatar*_are respectively the phase of the movement of the participant and the VP, both extracted from a Hilbert transform, | |is the norm in the complex plane, and *N* is the total number of samples;

iii/ a task score which is based on the ongoing relative phase of the VP and the participant’ s one, taking into account the task condition.

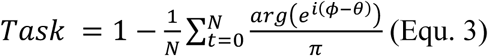

where *ϕ* is the relative phase, *i*.*e*. the difference between the phase of the movement of the participant and the VP, *θ* is the goal of the task, i.e. 0 if in-phase and *π* if anti-phase, N is the total number of samples, and arg() indicate the mathematical function returning the angle in radians of a complex number;

iv/ an intention score which evaluates the ability for the participant to properly attribute intention towards the “cooperative” or “competitive” behaviour of the VP.

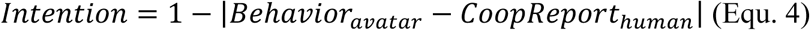

where *Behavior*_*avatar*_is 0 when the VP is competitive *i*.*e*. conflicting goal with the participant, and 1 when it is cooperative (i.e. shared goal with the human participant; and *CoopReport*_*human*_ is the rating of cooperativeness of the avatar reported by the human participant at the end of the trial;

and v/ a humanness score which reflects quantitatively the impression of the participant on the human or robotic character of the partner.

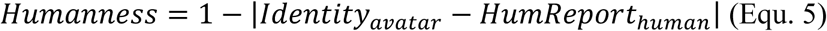

where *Identity*_*avatar*_is 0 when the VP is a robot and 1 when it is a human, and *HumReport*_*human*_ is the rating of humanness of the VP reported by the participant at the end of each trial.

### Statistical analysis and Normative Modelling

All statistical data analyses were performed using the Python 3.7 (Millman et Aivazis 2011) (numpy 1.17.2 (Oliphant 2006) and scipy 1.3.1 (Virtanen et al. 2019)): differences between groups, multiple linear regression models, and normative modelling. The normative modelling (NM) provides a metric similar to a Z-score, but accounts for the underlying structure of the population across multiple covariates. NM uses Gaussian Processes (GP) to model the distribution of control group measures while estimating separately the overall trajectory in the covariate space, the heterogeneity in the population, and the uncertainty of the fit (Zabihi et al. 2019). The Python code is available at https://github.com/GHFC/SoNeTAA/.

## 3. RESULTS

### 3.1 Socio-demographic and group comparative analyses

Children with ASD were younger than the control group (t(154)=2.6, p=2.6e-11**)), had a larger male/female ratio (Fisher exact, p=0.002), and scored higher in the SRS (t(154)=14.3, p=7.4e-29**). No statistically significant differences were found for IQ and handedness.

All the standardized psychometric instruments used showed significantly lower scores in the ASD group compared to the control groups: NEPSY-II TOM total score (Mann-Whitney U=104.5, p=0.0005**), NEPSY-II AF raw (U=114, p<0.002**), Purdue Pegboard score (U=138, p<0.0006**).

### 3.2 Correlation of chronological age with HDC scores

In a simple correlation analysis, task comprehension (r=0.33; p<0.005**) (Fig.2A), intention attribution (r=0.3; p<0.005**) (Fig.2B) and humanness (r=0.27; p<0.005**) (Fig.2C) scores showed substantive positive correlations with the chronological age in the entire cohort. A significant interaction was observed between the chronological age and the task comprehension score in the control group (r=0.4; p<0.05*) (Fig.2A), and with the intention attribution (r=0.21; p<0.5*) and the humanness (r=0.38; p<0.005**) scores in the ASDs group.

### 3.3 Comparison with standardized tests

Using normative modeling we virtually corrected for the developmental bias on the HDC scores. We then tried to observe how those corrected scores related to standard neuropsychological tests (Table 2).

**Table 2.** Summary of the main correlations between HDC scores and those from the NEPSY-II (Affect Recognition (AF) & Theory of Mind (TOM) Subdomains), the SRS-2 Social Responsiveness Scale - second edition and the Purdue Pegboard.

|  | Motor (NM) | Coordination (NM)<br>(NM) | Task (NM) | Intention (NM) | Humanness |
| --- | --- | --- | --- | --- | --- |
| SRS-2<br>p=0.6 | r=-0.22; p=0.01* | r=0.0031; p=0.97 | r=-0.22; p=0.0086* | r=-0.15; p=0.076 | r=-0.045; |
| NEPSY-II TOM<br>p=0.55 | r=-0.081; p=0.59 | r=-0.35; p=0.016* | r=-0.26; p=0.08 | r=0.17; p=0.24 | r=-0.09; |
| NEPSY-II AF<br>p=0.48 | r=0.2; p=0.18 | r=-0.043; p=0.78 | r=0.33; p=0.023* | r=0.16; p=0.3 | r=0.11; |
| Purdue Pegboard<br>p=0.34 | r=0.14; p=0.31 | r=-0.2; p=0.16 | r=-0.015; p=0.91 | r=-0.25; p=0.07 | r=0.13; |

We observed a significant interaction effect between the SRS-2 and motor score (r=-0.22; p<0.05*) (Fig.3A). NEPSY-II test showed significant interaction effect between its AF score and the HDC task comprehension score (r=0.33 p<0.05*) (Fig.3B). Supplementary data contain the details of the correlation per group, and with the HDC scores before the normative modeling correction.

### 3.4 Global comparative analysis between the controls and the ASDs groups

The comparative analysis between the two groups shows a statistically significant decrease in the ASDs group for the motor score (d=-0.5; p<0.005**). We also observed a better understanding of the task among ASDs participants (d=0.23; p<0.05*). Interactions between the two groups for the other scores (Coordination: d=-0.21, p=0.12; Intention: d=-0.12, p=0.49; Humanness: d=0.12, p=0.19) were not significant (Fig.4).

### 3.5 HDC scores analysis by sub conditions

HDC paradigm comes with sub conditions (see Figure 5 for a summary). Multiple regressions were thus calculated to predict the different normalized HDC scores based on the diagnostic (coded as 0 = ASD and 1 = CTR), the age (in years), the avatar behavior (coded as 0 = Competitive and 1 = Cooperative), and the human task (coded as 0 = Anti-phase and 1 = In-phase).

**Figure 4.**
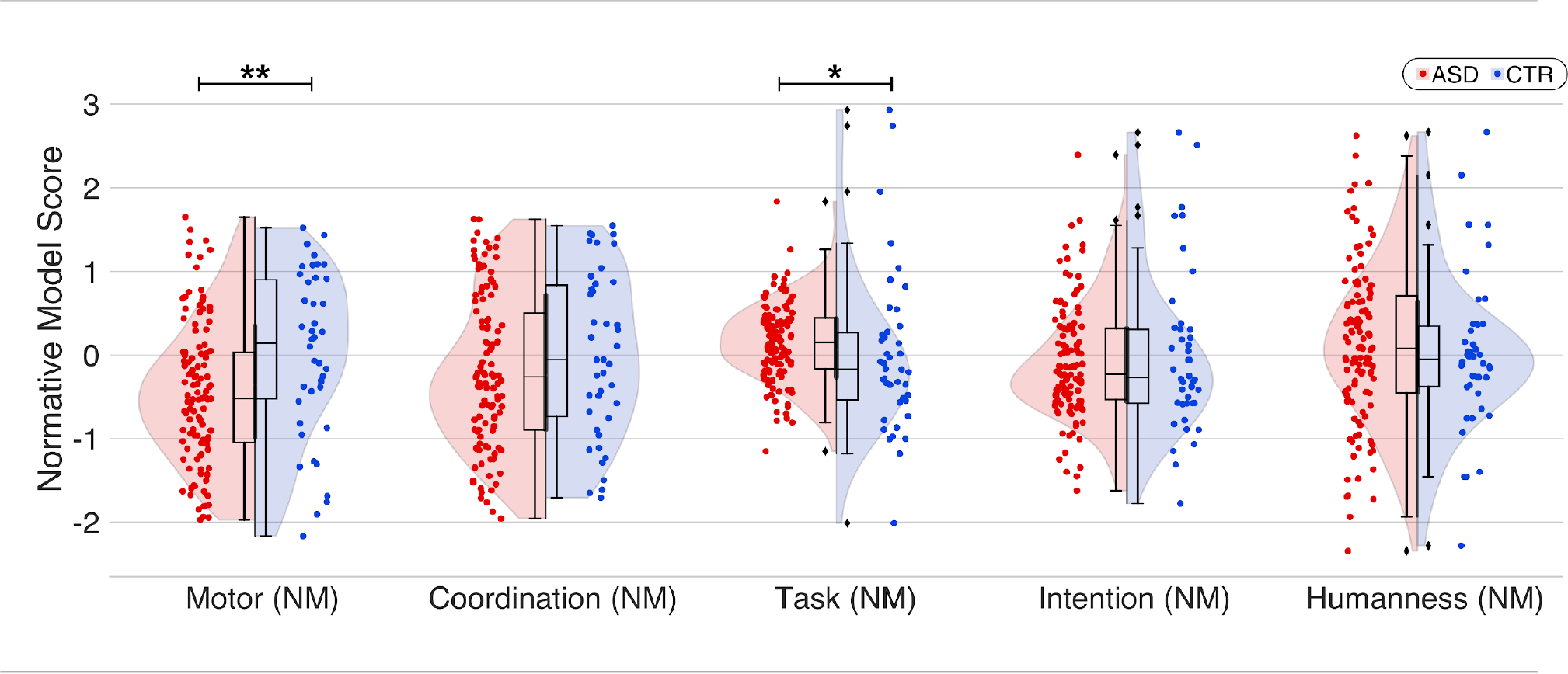
Comparison between the ASDs and control groups across different behavioral scores derived from the HDC protocol and corrected with Normative Modeling. Only motor score really discriminates the two population (d = −0.5; p<0.005**), with significant lower results among ASDs. The lines represent linear regressions. Coloured areas: 95% confidence intervals (CI). Neurotypical participants: blue, ASDs: red.

**Figure 5.**
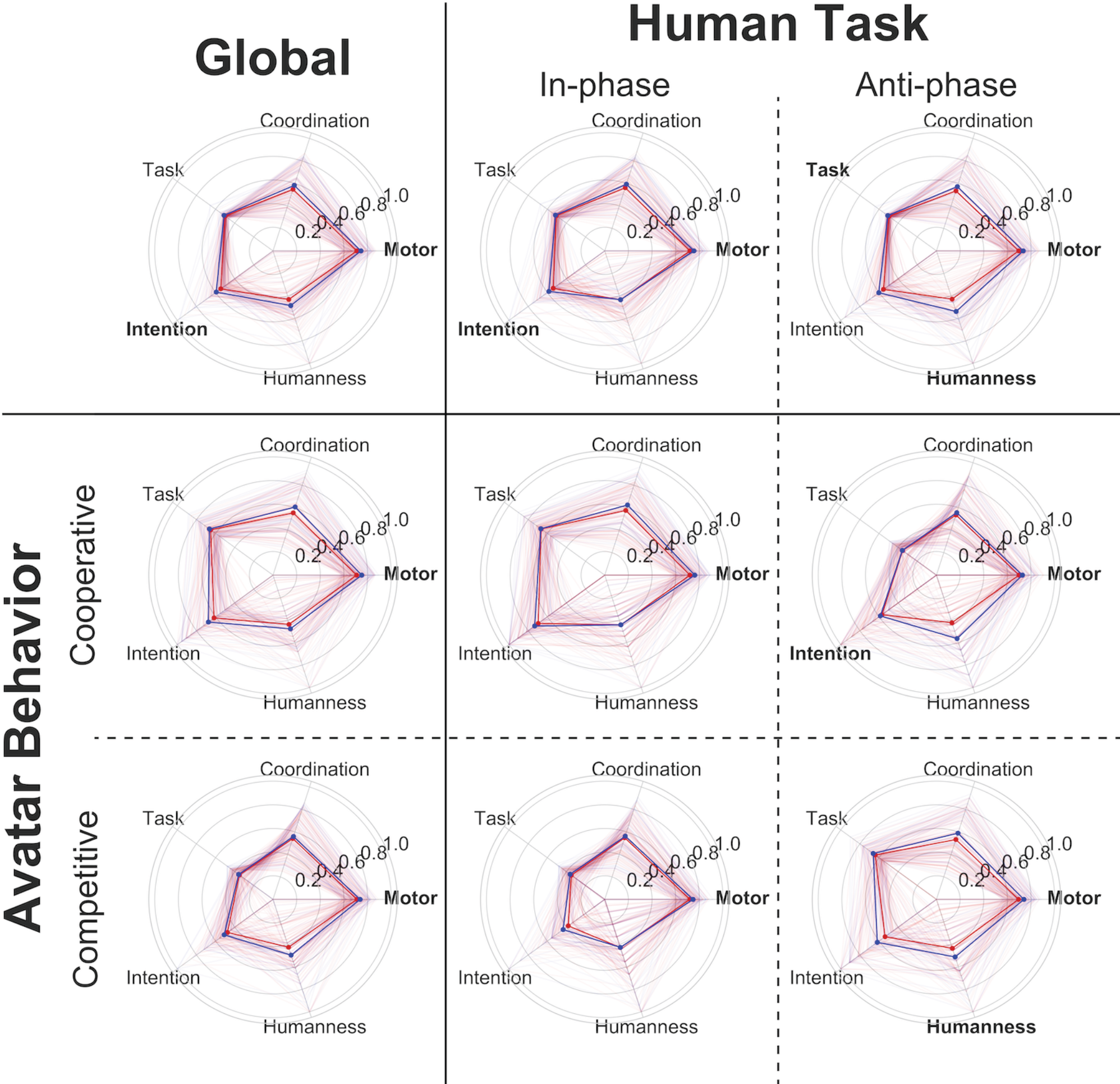
Score analysis by sub-conditions (Human task: in-phase/anti-phase and Avatar behavior: cooperation/competition), in the two groups. Both the diagnostic (see Global) and the Human Task were significant predictors of the motor score, with Control group having greater scores (coeff = 0.4413, p < 0.001**), and “in-phase” task leading as well to better scores (coeff = 0.2726, p = 0.016*). Controls are in blue and ASDs participants in red. Scores with a statistically significant differences between ASDs and Controls are in bold typeface.

There was a significant regression equation for the motor score (F(5, 618) = 7.634, p = 5.64e-07). Both the diagnostic and the human task were significant predictors of the motor score, with Control group having greater scores (coeff = 0.4413, p < 0.001), and in-phase task leading as well to better scores (coeff = 0.2726, p = 0.016). There was a significant regression equation for the Coordination score [F(5, 618) = 3.252, p = 0.00659]. Age was a significant predictor of the Coordination score, with a positive effect of age on coordination (coeff = 0.0272, p = 0.022). There was a significant regression equation for the Task score [F(5, 618) = 409.1, p = 2.42e-193]. Avatar behavior was a significant predictor of the Task score, with a cooperative behavior of the VP having a huge effect on the task comprehension in the participant (coeff = 6.9602, p < 0.005). There was also a significant regression equation for the Intention score [F(5, 618) = 28.84, p = 2.46e-26]. Both Human task and Avatar behavior were significant predictors of the Intention score. Participants tend to better detect the intention of the VP while “anti-phase” (coeff = −1.2718, p < 0.005), and while “in-phase” if the VP takes a cooperative behavior (coeff = 2.3107, p < 0.005).

For Motor score, detailed analysis suggests that during the VP “cooperation” behavior, the task allows to distinguish the two groups “in-phase”: d=−0.51; p=0.0059* and “anti-phase”: d=−0.59; p=0.0015**. Furthermore, during the VP “competitive” behavior, the task allows to distinguish the two groups “in-phase”: d=−0.34; p=0;025* and “anti-phase”: d=−0.35; p=0.025* (Table 3).

**Table 3.** HDC scores analysis by sub conditions.

|  | Motor (NM) |  | Coordination (NM) |  | Task (NM) |  | Intention (NM) |  | Humanness (NM) |  |
| --- | --- | --- | --- | --- | --- | --- | --- | --- | --- | --- |
|  | coop | comp | coop | comp | coop | comp | coop | comp | coop | comp |
| in-phase | d=-0.51<br>(p=0.0059*) | d=-0.34<br>(p=0.025*) | d=-0.26<br>(p=0.098) | d=-0.087<br>(p=0.35) | d=0.12<br>(p=0.17) | d=0.067<br>(p=0.47) | d=-0.018<br>(p=0.47) | d=-0.084<br>(p=0.35) | d=0.34<br>(p=0.014*) | d=0.3<br>(p=0.046*) |
| anti-phase | d=-0.59<br>(p=0.0015**) | d=-0.35<br>(p=0.025*) | d=-0.3<br>(p=0.054) | d=-0.11<br>(p=0.29) | d=-0.067<br>(p=0.41) | d=0.16<br>(p=0.23) | d=-0.2<br>(p=0.14) | d=0.067<br>(p=0.31) | d=0.012<br>(p=0.49) | d=-0.25<br>(p=0.058) |

Coop = cooperative behavior of the VP , comp = competitive behavior of the VP, d = cohen-d, p = p-value, \* = $p < 0.05$ , \*\* = $p < 0.005$ .

## 4. DISCUSSION

### 4.1 Interpretations of the results

#### Developmental aspects of socio-cognitive skills and intervention based on IS in ASDs children

Literature about IS attest of its developmental aspect and plasticity allowing therapeutic detections and actions (Landa et al. 2013; 2011; Franz et Dawson 2019). Our results are in line with the developmental aspect of socio-cognitive skills (intention attribution: r=0.21; p<0.05*; humanness: r=0.3; p<0.005** and task comprehension: r=0.4; p<0.05*). To go further, interventions targeting early development of socially synchronous interaction in toddlers with ASDs have demonstrated its effectiveness (Landa et al. 2011), suggesting improvement in child language comprehension and severity of the ASDs symptoms (Tachibana et al. 2017).

#### Coupling between low-level sensory-motor, motor and high-level socio-cognitive skills

Better affect recognition was associated with a better comprehension (r=0.33; p=0.023*). One hypothesis that could be formulated would be that of a mediating effect of IQ on the recognition of emotions, suggesting that greater cognitive abilities correlates with a higher levels of affect recognition skills. Results attest of lower motor skills among participants with higher probability of dimensional diagnosis of ASDs (SRS-2: r=-0.22; p=0.01*). Motricity in ASDs will be discussed further, but a first conclusion of this results suggests a substantial link between low-level motor skills and high-level social-cognitive skills among a population of children and adolescents. Some support a strong pairing between the mirror system and the mentalizing system during communicative gestures, suggesting a cognitive-motor coupling in children (Nadel 2015). The mechanisms involved are not only mechanistic but are also part of neurobiological processes, characterized by the release of endogenous opioids (dopamine, endorphins, serotonin and oxytocin) (Mu et al. 2016; Lang et al. 2017) and the recruitment of now well described neural basis (Bhat et al. 2017). Multiple mechanisms are actually debated to explain synchrony in an evolutionary perspective. IS plays a role in shared common goals that lead to cooperative expectations and joint action behaviors (Valdesolo et al. 2010), in shared basic affective states and emotion, in a better attribution of his one’ s self and others and in a better comprehension of the social situations (Valdesolo et DeSteno 2011).

#### Motor skills as a developmental marker of children and adolescents at risk with ASDs

Motor score is the only HDC scores that allows to distinguish the two groups (coeff = 0.4413, p < 0.001**). In overall terms, this motor low-level score is found to be statistically lower among ASDs participants, confirming current data finding altered motor skills in ASDs. Alterations in motor control (Fournier et al. 2010), and particularly of the executive motor control (Demetriou et al. 2018) have been widely demonstrated in ASDs children. However, while motor disorders are associated with the diagnosis of ASDs children in 50 to 80% (Green et al. 2009) and that their prevalence increased with age (Licari et al. 2019), they remain under-diagnosed in clinical practice (1.34%) (Licari et al. 2019). On the other hand, their estimated prevalence (36%) makes them almost as frequent as cognitive disorders (38%) among children under 6 (Licari et al. 2019). Thus, ASDs children and adolescents tend to have difficulties in planning and sequencing their movements (Grace et al. 2018) and are associated with higher levels of neuromotor noise (Torres et al. 2017) (*i*.*e*. disturbing action (motor commands) and perception (sensory feedback) (Xavier et al. 2018)). This noise can have multiple substrates but relies on hypotheses which can be explained with Bayesian models with imbalance between prediction, inputs and expectations within sensory-motor integration (Bolis et Schilbach 2017). Indeed, ASDs are associated with alterations of the integration of social stimuli (Thye et al. 2018) and a reduced ability to integrate somatosensory and visual information into accurate motor responses (Lim et al. 2017). We also formulate different hypotheses to explain these results. Children with ASDs could have difficulties to maintain their attention for a time long enough to perceive the stimulus with satisfactory attention. Though, some studies now described deficit of joint-attention as an endophenotype of ASDs (Mundy et al. 2017; Jones et al. 2017). Further analysis showed that the instruction given to the participant (Human task: “in-phase”) is associated with a better motor score among the control group (coeff = 0.2726, p = 0.016*). Furthermore, we observed interactions by sub condition. ASDs children tend to have lower results “in-phase” while a “competitive” and cooperative” behavior of the avatar and during “anti-phase” while a “competitive” and cooperative” behavior of the avatar (Table 3). Thus, Wang and colleagues (2019) reported that during a cooperative task of synchronization (i.e. “in-phase”), children with severe diagnostic of ASDs tend to have lower action level.

#### Socio-cognitive skills based on Interpersonal Synchrony

Mentalizing deficits have repeatedly been described in ASDs population (Bliksted et al. 2016). This process need preserved metacognitive skills, yet metacognitive monitoring is found diminished in children with ASDs (Grainger et al. 2016). Higher-level scores (intention attribution and task comprehension) require better use of metacognitive processes. However, for both score we did not observe an effect of the group. Thus, for the intention attribution score, it seems easier to detect the intention of the VP while “anti-phase” (coeff = −1.2718, p < 0.005), and while “in-phase” if the VP takes a cooperative behavior (coeff = 2.3107, p < 0.005). Furthermore, the comprehension of the task is better if the Avatar is cooperative (coeff = 6.9602, p < 0.005). One result has caught our attention and seems difficult to interpret. ASD participants would have a better understanding of the task than controls (d=0.23; p<0.05*). One hypothesis that could be made is that ASDs participants tend to generate mainly the same movement “in-phase” with the avatar without taking the instruction into account. This bias could be responsible for a false positive result.

### 4.2 Implications findings for clinical practice and public health

The aim of this study is also to give standardized values to assess the HDC. We generated percentile ranks for each HDC score from the results obtained in the control patients (see Supplementary Table 1). In this way it is possible to estimate a child’ s skills for each assessment.

Research has tried to identify clinical markers in ASDs: early patterns (Ozonoff et al. 2018), a regression phenomenon (Ozonoff et Iosif 2019), an atypical neural response of the gaze (Jones et al. 2014). Later, neurological soft signs (Tani et al. 2006), abnormalities of sensorimotor priors (Torres et al. 2013), anomalies in proprioceptive and sensory-motor development (alteration of motor priors, micromovements and the presence of noise in sensory-motor variables causing a lack of embodiment) (Torres et al. 2013). Systematic and reliable metrics of the HDC, normalized across developmental trajectories through normative models (Lefebvre et al. 2018), could allow to predict phenotypic profiles, and thus refine the diagnosis, associated comorbidities and the study of the disorder through stratification.

## Conclusion

The HDC can evaluate interpersonal synchrony at both the low and high level of social cognition during live interaction, but also probe the developmental aspects of their relationship. On the other hand, the psychometric evaluation of HDC provides reliable, reproducible, objective, standardized scores, derived from a natural movement. As a new psychometric test, it could identify motor and social markers to improve early detection of neurobehavioral abnormalities during human interaction. This paradigm also provides a basis for the development of therapeutic approaches, for example the serious game “Pop’ Balloons” that allows an automatic evaluation in mixed reality (demonstration video: https://vimeo.com/277085489).

## Data Availability

The datasets generated during and/or analyzed during the current study are not publicly available due to restriction of ethical consents but aggregated data are available from the corresponding author on reasonable request.

## AUTHOR CONTRIBUTIONS

FB, AL, AP collected all the data. FB, AL, FA, and AM worked on the inclusion of patients and their clinical exploration. FB, AL, YB, DE participated in both analysis and writing. RD, and TB participated in the design and relecture. GD participated in the design, analysis, and writing.

## ACKNOWLEDGMENTS

The authors would like to thank the participants and their families who participated in this study. They also thank Benjamin Landman, Anna Banki, and Ramon Aparicio-Garcia for early help in the project.

## SUPPLEMENTARY MATERIAL

**Supplementary Table 1.** HDC percentile ranks by age group.

| Age | 5-10 |  |  | 10-13 |  |  | 13-15 |  |  | 15-26 |  |  |
| --- | --- | --- | --- | --- | --- | --- | --- | --- | --- | --- | --- | --- |
| Percentiles | 5e | 50e | 95e | 5e | 50e | 95e | 5e | 50e | 95e | 5e | 50e | 95e |
| Motor | 0.65 | 0.72 | 0.86 | 0.63 | 0.70 | 0.81 | 0.62 | 0.79 | 0.88 | 0.58 | 0.73 | 0.88 |
| Coordination | 0.24 | 0.55 | 0.85 | 0.24 | 0.40 | 0.83 | 0.32 | 0.63 | 0.90 | 0.325 | 0.54 | 0.89 |
| Task | 0.23 | 0.48 | 0.80 | 0.27 | 0.50 | 0.75 | 0.19 | 0.49 | 0.79 | 0.24 | 0.50 | 0.84 |
| Intention | 0.26 | 0.51 | 0.69 | 0.34 | 0.56 | 0.87 | 0.0 | 0.61 | 1.0 | 0.18 | 0.58 | 1.0 |
| Humanness | 0.0 | 0.17 | 0.30 | 0.27 | 0.50 | 0.76 | 0.07 | 0.55 | 1.00 | 0.0 | 0.50 | 0.95 |

